# Robust Discovery of Mild Cognitive impairment subtypes and their Risk of Alzheimer’s Disease conversion using unsupervised machine learning and Gaussian Mixture Modeling

**DOI:** 10.1101/2020.10.14.20212696

**Authors:** Fahimeh Nezhadmoghadam, Antonio Martinez-Torteya, Victor Treviño, Emmanuel Martínez, Alejandro Santos, Jose Tamez-Peña, for the Alzheimer’s Disease Neuroimaging Initiative

## Abstract

**Background:** Alzheimer’s disease (AD) is an irreversible, progressive brain disorder that slowly destroys memory and thinking skills. The ability to correctly predict the diagnosis of Alzheimer’s disease in its earliest stages can help physicians make more informed clinical decisions on therapy plans.

**Objective:** To determine whether the unsupervised discovering of latent classes of subjects with mild cognitive impairment (MCI) may be useful in finding different prodromal AD stages and/or subjects that have a low MCI to AD conversion risk.

**Methods:** 18 features relevant with the MCI to AD conversion process described 681 subjects with early MCI. Subjects were split into training (70%) and validation (30%) sets. Subjects from the training set were analyzed using consensus clustering and Gaussian mixture models (GMM) were used to describe the shape of the discovered latent classes. The discovered GMM predicted the latent class of the validation set. Finally, descriptive statistics, rates of conversion, and odds ratios (OR) were computed for each discovered class.

**Results:** Through consensus clustering we discovered three different clusters among MCI subjects. The three clusters were associated with low-risk (OR = 0.12, 95%CI = 0.04 to 0.3|), medium-risk (OR = 1.33, 95%CI = 0.75 to 2.37), and high-risk (OR = 3.02, 95%CI = 1.64 to 5.57) of converting from MCI to AD, with the high-risk and low-risk groups highly contrasting. Hence, prodromal AD subjects were present on only two clusters.

**Conclusion:** We successfully discovered three different latent classes among MCI subjects with varied risk of MCI-to-AD conversion through consensus clustering. Two of the discovered classes may represent two different prodromal presentations of the Alzheimer’s disease.

## 1. INTRODUCTION

Alzheimer’s disease (AD) is the most common form of dementia among the elderly [1]. There are currently more than 50 million people worldwide with dementia and 131.5 million people are predicted to be affected by AD in the year 2050 [2]. Hence there is a need for the discovery of effective treatments aimed to improve or cure the future AD cases. The clinical research and the pharmaceutical community have explored numerous new drugs, however over the last twenty years, no effective treatments have been developed to aid the affected population [3]. The current hypotheses for the lack of success in AD drug development are that current drugs cannot change the course of dementia at late disease stages and that subtle differences in AD phenotypes may require different treatment. Therefore, it is extremely important to diagnose AD in its earliest stages as well as detecting the possible AD subtype affecting the patient. Hence, early diagnosis and subtyping of Alzheimer’s disease have several benefits: They play a primal role in prompt evaluation and treatment of reversible or treatable causes and help physicians to make important decisions about patient’s care [4].

To achieve the early detection goal it is important to get a clear understanding of the causes and pathogenesis of AD and differentiate it from the complexity of aging [5-7]. It has been reported that 33.6% of subjects with mild cognitive impairment (MCI) are at risk of having early-stage AD [8, 9]. Hence, the prodromal phase of the AD is characterized by MCI with AD pathological features in brain tissue that start early in the disease process and are considered hallmarks of the disease [10]. In other words, the MCI stage in AD patients is a transitional stage between normal aging and clinical dementia [8, 9].

Some of the reported diagnostic methods are not accurate enough to separate patients with MCI that will progress towards AD *vs* subjects that will develop another type of dementia [11-13]. Therefore, there is a demand for developing precise prediction models that yield an adequate prediction of subjects that will develop AD. Various methods have been suggested for early diagnosis [2, 14-16]. Although some work has been done in identifying different presentations of AD at an early stage [17], these efforts have been done in different settings and using limited clinical information [18]. The main challenge of AD subtyping is that the prodromal phase is defined by a collection of symptoms that are shared by many diseases or conditions [19]. Furthermore, there is a wide variety of possible disease subtypes and each one is associated with the explicit clinical data type used to define it: Cognitive questionnaires, medical history, imaging features, laboratory tests, nutrition habits, genomics, proteomics, radiomics, and so forth [20]. Hence, reaching a consensus definition of clinically relevant disease subtypes at the prodromal stage is of paramount importance [21, 22].

Defining clinical relevant subtypes is challenging, because there are thousands of possible ways in which MCI patients could be classified, and there is no clear difference in survival trajectories between many MCI to AD converters. One of the suggested methods proposes to apply a novel multi-layer clustering algorithm to a longitudinal cohort of MCI subjects to identify homogenous subtypes [23], while Ezzati et al. (2019) applied a latent class analysis (LCA) to detect subgroups among MCI patients [18]. To address this issue, we propose to use robust and unsupervised data clustering of the multidimensional features of MCI patients [24]. Data clustering aims to find a computer model that is able to select subjects that share similar characteristics. Our hypothesis is that those robust clusters also share the same AD etiology, thus, they may have similar treatment responses. Although data clustering can be done by many different machine learning (ML) algorithms, recent advances in ML have set forward statistical clustering strategies that are robust to algorithmic approaches [25, 26]. One of the most robust approaches applies consensus clustering, and Gaussian mixture models (GMM) to discover the stable latent classes from multidimensional data sets [27, 28]. The objective of this study was twofold: To apply consensus data clustering to discover the latent classes from a set of AD-relevant characteristics from MCI subjects, and to describe the association of discovered classes with MCI to AD conversion.

## 2. MATERIALS AND METHODS

### 2.1. Data

Data used in the preparation of this study was obtained from the TADPOLE challenge “standard” data sets (https://tadpole.grand-challenge.org). The TADPOLE sets were derived from the Alzheimer’s Disease Neuroimaging Initiative (ADNI) study (adni.loni.usc.edu). The ADNI was launched in 2003 by the National Institute on Aging, the National Institute of Biomedical Imaging and Bioengineering, the Food and Drug Administration, private pharmaceutical companies and non-profit organizations, as a $60 million, 5-year public-private partnership. The primary goal of ADNI has been to test whether serial MRI, positron emission tomography (PET), other biological markers, and clinical and neuropsychological assessment can be combined to measure the progression of MCI and early AD. Determination of sensitive and specific markers of very early AD progression is intended to aid researchers and clinicians develop new treatments and monitor their effectiveness, as well as lessen the time and cost of clinical trials. The Principal Investigator of this initiative is Michael W. Weiner, MD, VA Medical Center and the University of California – San Francisco. ADNI is the result of efforts of many co-investigators from a broad range of academic institutions and private corporations, and subjects have been recruited from over 50 sites across the U.S. and Canada. The initial goal of ADNI was to recruit 800 subjects but ADNI has been followed by ADNI-GO and ADNI-2. To date, these three protocols have recruited over 1500 adults, ages 55 to 90, to participate in the research, consisting of cognitively normal older individuals, people with early or late MCI, and people with early AD. The follow-up duration of each group is specified in the protocols for ADNI-1, ADNI-2, and ADNI-GO. Subjects originally recruited for ADNI-1 and ADNI-GO had the option to be followed in ADNI-2. For up-to-date information, see www.adni-info.org.

The ADNI-TADPOLE dataset included 1737 subjects with longitudinal observations. Each observation included the diagnosis status, neurocognitive evaluations, quantitative MRI longitudinal observations, PET studies, APOE4 polymorphism, among others [29]. For this study, only the 681 subjects with early MCI (EMCI) and late MCI (LMCI) at baseline were included. Indeed, the ADNI dataset separates “early” and “late” mild cognitive impairment (MCI) based on a single memory test so that EMCI subjects are a cohort with milder episodic memory impairment than the LMCI group [30, 31]. This dataset comprised the different clinical data types: Questioners, Radiomics-Imaging, Laboratory test, Genomic.

### 2.2. Feature selection

Although the ADNIT-TADPOLE data set described each subject with 1,898 features, we only used a basic set of 18 features that can readily be observed in clinical practice and that are relevant or associated with MCI to AD conversion. Selected features included demographic information, scores from cognitive tests, volumes from different brain regions as measured through MRI, and genetic information. Information derived from PET imaging and cerebrospinal fluid were not considered due to the difficulty of performing those analyses across the world.

The three selected demographic features were: Age, gender, and years of education. Nine features from cognitive tests were selected, namely: The 11-item score and 13-item score from the Alzheimer’s Disease Assessment Scale (ADAS); total score from the Clinical Dementia Rating scale Sum of Boxes (CDR-SB); total score from the Functional Assessment Questionnaire (FAQ); total score from the Mini-Mental State Examination (MMSE); and immediate score, learning score, forgetting score, and percentage forgetting score from the Rey’s Auditory Verbal Learning Test (RAVLT). The five MRI-related features were: Total entorhinal volume, hippocampal volume (left plus right), middle temporal gyral volume (left plus right), total ventricular volume, and intracranial volume (ICV). Finally, the data set also included APOE4 status.

The rationale for some of the selected features is that several studies have established some association between them and the AD neurodegenerative process [32]: Cognitive evaluations have strong predictive power of MCI to AD conversion, hippocampal and entorhinal volumes have also been associated with the MCI to AD conversion process, and APOE4 has been shown to be associated with age at onset of AD [33-38]. The demographic features, ventricle volume, and ICV were selected to study any possible anthropometric-, age-, or education-related associations in the clusters. Finally, the time to AD conversion was computed to study discovered MCI subtypes, while early MCI (EMCI) and late MCI (LMCI) status were selected to describe the associations of the discovered clusters.

Selected features at baseline are shown in Table 1. While the sex and age of both classes were slightly similar, there was a statistical difference between the ratio of MCI to AD converters. Finally, we computed the effect size of all features using Cohen’s d and odds ratio for continuous and discrete variables, respectively [39].

**Table 1.** Baseline characteristics of Tadpole Challenge Subjects used on this study.

| Feature | EMCI | LMCI | Effect Size |
| --- | --- | --- | --- |
| Subjects (male ratio) | 249 (55.02%) | 432 (61.11%) | OR=1.28 (0.94-1.76) |
| Age | 70.98 (0.46) | 73.31 (0.36) | Z=0.32* |
| Years of Education | 16.03 (2.65) | 15.77 (3.01) | Z=0.08* |
| MMSE total score | 28.37 (0.1) | 27.15 (0.09) | Z=0.72** |
| ADAS13 | 12.54 (0.33) | 18.59 (0.31) | Z=1.02*** |
| ADAS11 | 7.84 (0.22) | 11.49 (0.22) | Z=0.90*** |
| CDR-SB total score | 1.31 (0.05) | 1.64 (0.04) | Z=0.39* |
| RAVLT immediate score | 39.42 (0.67) | 31.42 (0.46) | Z=0.81*** |
| RAVLT learning score | 5.35 (0.15) | 3.44 (0.12) | Z=0.78** |
| RAVLT % forgetting score | 47.29 (1.94) | 68.31 (1.5) | Z=0.68** |
| RAVLT forgetting score | 4.33 (0.17) | 4.79 (0.11) | Z=0.19* |
| FAQ total score | 2.10 (0.21) | 3.89 (0.22) | Z=0.46* |
| Entorhinal cortex volume | 3.77E3 (42.13) | 3.33E3 (35.7) | Z=0.62** |
| Hippocampal volume | 7.25E3 (65.09) | 6.48E3 (53.13) | Z=0.72** |
| Middle temporal gyral volume | 20.75E3 (164.4) | 18.97E3 (142.09) | Z=0.64** |
| Ventricular volume | 34.93E3 (1.31E3) | 42.67E3 (1.14E3) | Z=0.35* |
| ICV | 1.51E6 (9.74E3) | 1.56E6 (8.03E3) | Z=0.29* |
| APOE4: {0, 1, or 2} ε4 alleles | {145, 90, 14} | {192, 177, 63} | OR=0.57 (0.42- 0.79) |
| MCI to AD converter ratio | 11.65% | 49.77% | OR=7.5 (4.9- 11.5)*** |

Values in parenthesis represent the mean standard error unless specifically stated. The OR was computed with a confidence interval of 95% for LMCI vs EMCI. \*, \*\*, and \*\*\* denote a small effect size (between 0.2 and 0.5 for Z and between 1.5 and 2 for OR), a medium effect size (between 0.5 and 0.8 for Z and between 2 and 3 for OR), and a large effect size (larger than 0.8 for Z and more than 3 for OR), respectively.

### 2.3. Consensus Clustering and Gaussian Mixture Modeling

Figure 1 summarizes the overall methodology used for cluster discovering and validation. The specific model parameters depend on the training set [40], hence, model fitting and inference are biased towards the training set. To avoid training biases, our set of MCI subjects was split into training and validation sets: 70% (n=476) of the subjects were randomly selected and used to discover the MCI clusters/latent classes and to train the GMM parameters. Once all the cluster-parameters were estimated on the training set, we predicted the corresponding classes on the remaining 30% (n=205) of the subjects. After that, we described the characteristics of each one of the groups.

**Figure 1.**
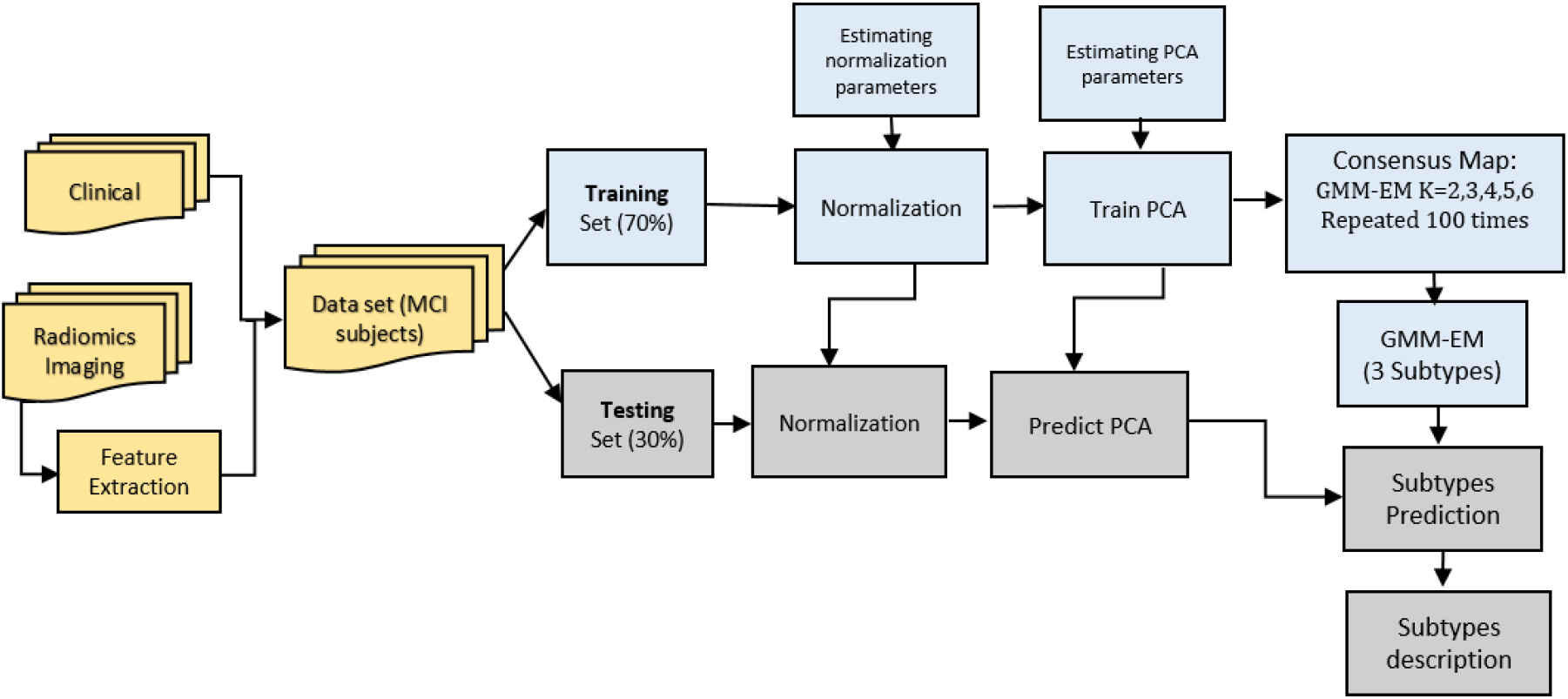
The overall methodology of subtype’s classification of ADNI dataset. The multimodal data is split into training and testing sets and the results of the testing set are used to describe the association of disease subtypes to clinical relevant outcomes.

First, we had to estimate the optimal number of *K* Gaussian mixtures and parameters *θ =* {*ϕ*_1_, *μ*_1_, *Σ*_1_, …, *ϕ*_*K*_, *μ*_*K*_, *Σ*_*K*_} given the multidimensional vector *x* of each patient in the training set. In other words, the algorithm must find the optimal partition of the data (*K*) and the parameters of the Posterior probability distribution described by eq. 1:

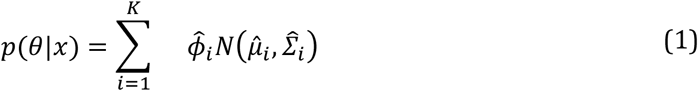

where 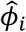 is the mixture weight, and 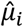 and 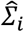 are the mean vector and covariance matrix of each Gaussian cluster, respectively [41]. Once a GMM was estimated, the latent class *ŷ* of a new subject was then predicted using the Mahalanobis distance (eq. 2):

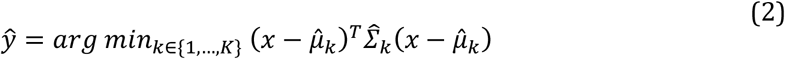

Not all the observed features follow a Gaussian distribution, but we assumed that the linear combination of random variables, eq. 3:

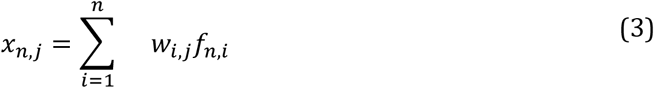

follows a normal distribution, where *w*_*i,j*_ is a coefficient that weights the contribution of each *f*_*i*_ feature for a specific patient *n*. In this study, we used the principal components analysis (PCA) transform to find the weights of the linear transformation [42-44]. To avoid feature biases in the PCA computation, we normalized all the features using the z-transform:

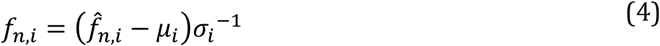

where 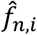 is the raw observed feature, and *μ*_*i*_ and *σ*_*i*_ are the sample mean and the sample standard deviation of each feature, respectively.

We estimated *K* using the consensus clustering method, a robust methodology for the estimation of the clustering structure of the sampled data set [45-48]. The main element of consensus clustering is the cluster co-association matrix (CCAM), an NxN matrix that stores the frequency that two subjects share the same latent class. The frequency is computed by randomly selecting a training set of subjects and then estimating their latent class by a user-supplied clustering method. After that, we count if a subject pair shares the same cluster, e.g.:

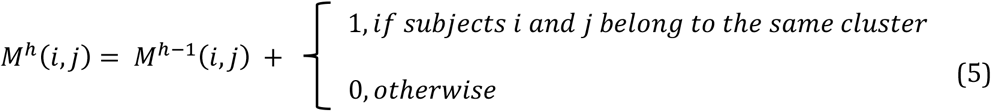

where *M*^*h*^ is the CCAM after the *h*^*th*^ repetition of the clustering method. The procedure is repeated several times to get a robust estimation of the co-association. The analysis of the CCAM matrix is done by computing the proportion of ambiguous clustering (PAC) and studying the hierarchical cluster of the CCAM [27].

In summary, the specific implementation of the consensus clustering was as follows: First, we normalized all features; second, we performed the PCA transform and selected the first 3 components (capturing more than 80% of total variance); third, we repeated the clustering method for *K*= {2, 3, 4, 5 *and* 6) one hundred times, where at each specific *K*, we randomly selected 70% of the training samples for latent class discovery using the expectation-maximization (EM) algorithm of the mclust package (mclust 5.4.5) [49] and the latent classes of the other 30% (i.e. the clustering validation set) were predicted using equation (2) with the trained GMM; fourth, the CCAM matrix of the predicted latent classes was analyzed and used to discover the optimal number of clusters; fifth, the GMM was fit and trained using the optimal number of clusters found by consensus clustering on the PCA transformed data using the EM algorithm (mclust 5.4.5).

### 2.4. Statistical Analysis of the MCI Subtypes

After we estimated the z-normalization parameters, computed the PCA transform, discovered the number of MCI clusters and their associated parameters of the GMM, we proceeded to estimate the latent class of each one of the samples in the holdout sample. The latent class prediction is done in three steps: First, compute the z-score of each one of the hold-out-samples using equation (4) and the mean and standard deviation of the train samples; second, extract the tree principal components using equation (3); and third, estimate the latent class using equation (2). This procedure provides a unique class label for each testing-set sample. Hence, to test the hypothesis that each latent class represents a unique set of clinically relevant MCI subtypes, we studied the association of each cluster to the clinical relevant outcome of MCI to AD conversion.

The relevance of each latent class in the context of AD was determined by computing the OR for MCI to AD conversion [50] and the Kaplan-Meier plot of each cluster [51], checking for statistical differences between the survival curves using the Log-rank test [52]. Furthermore, we studied the differences of each latent class among converters and non-converters (prodromal AD) and statistical differences between features of the subtypes were computed using the ANOVA test. Finally, we studied the potential training bias between the discovered subtypes by reporting statistical differences between training and testing sets [53]. Values lower than 0.05 were considered significant, and no attempt was made to correct for false discovery.

## 3. RESULTS

Through consensus clustering we discovered three different sets of MCI patients. Figure 2 shows the CCAM of the 100 predicted validation results of the consensus clustering for all analyzed values of *K*, and clearly illustrates these three different sets of patients among the ADNI-MCI. Additionally, Figure 2 also displays the result of the comparison of the PAC. Taking these results into account, we judged that *K* = 3 was optimal for this analysis.

**Figure 2.**
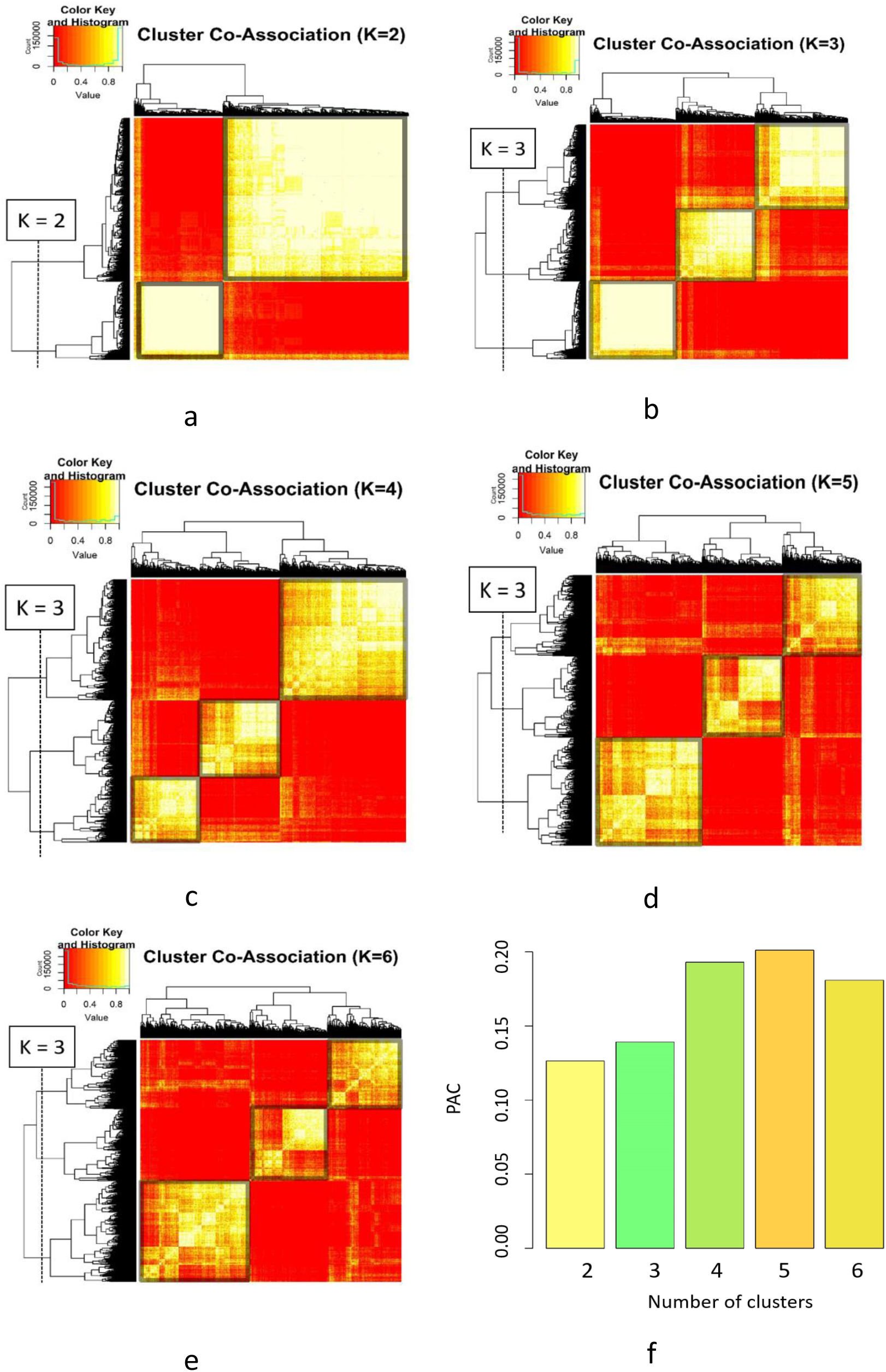
Results of consensus clustering applied to the validation set of MCI subjects of the ADNI dataset. a, b, c, d and e) Consensus mapping for *K* = 2,3,4,5 *and* 6, respectively, and f) the comparison of PAC (smaller is the better) between the cluster numbers from 2 to 6.

Figure 3 depicts the PCA and t-distributed stochastic neighbor embedding (t-SNE) visualization of the predicted latent class on the training set and the hold-out testing set. Subtypes 1 through 3 represent the MCI to AD low-, medium-, and high-risk subtypes, respectively. Table 2 illustrates the proportion of reverters (conversion from MCI to normal cognition), stable MCI subjects, and converters (conversion from MCI to AD) per discovered MCI subtype, as well as their corresponding OR with a 95% confidence interval. It is clear that, with an OR of 3.02 (95%CI 1.64 to 5.57), subtype 3 represents a group at a higher risk of MCI to AD conversion than the other two subtypes. On the other hand, subtype 1 has an almost null risk of converting, with and OR of 0.12 (95%CI 0.04 to 0.31). Hence, with an OR of 1.33 (95%CI 0.75 to 2.37), subtype 2 was considered a medium-risk subgroup. Furthermore, the low-risk subjects have a high chance of reverting back to a normal cognitive status, with an OR of 29.15 (95%CI 3.6 to 236.23). Additionally, the amount of EMCI patients in the medium- and high-risk groups was much smaller than LMCI subjects, whereas most subjects in the low-risk group were EMCI patients.

**Table 2.** The results of classification on testing set (N = 205).

|  | **Subtype 1**<br>**Low-Risk**<br>**(N=55)** | **Subtype 2**<br>**Medium-Risk**<br>**(N=85)** | **Subtype 3**<br>**High-Risk**<br>**(N=65)** | **Subtype 2 + Subtype 3**<br>**At-Risk**<br>**(N=150)** |
| --- | --- | --- | --- | --- |
| **EMCI** | 70.91% | 28.24% | 15.38% | 22.67% |
| **Reverters (MCI to NC)** | 16.36% | 0% | 1.54% | 0.67% |
| **Stable subjects** | 74.55% | 60% | 44.61% | 53.33% |
| **Converters (MCI to AD)** | 9.09% | 40% | 53.85% | 46% |
| **OR of reverting** | 29.15<br>(3.6 - 236.23) | - | 0.23<br>(0.03 - 1.83) | 0.03<br>(0 - 0.28) |
| **OR of remaining stable** | 2.56<br>(1.29 - 5.09) | 1.07<br>(0.61 - 1.89) | 0.42<br>(0.23 - 0.77) | 0.39<br>(0.2 - 0.78) |
| **OR of converting** | 0.12<br>(0.04 - 0.31) | 1.33<br>(0.75 - 2.37) | 3.02<br>(1.64 - 5.57) | 8.52<br>(3.22 - 22.56) |

The OR for each subtype were computed with a confidence interval of 95%.

**Figure 3.**
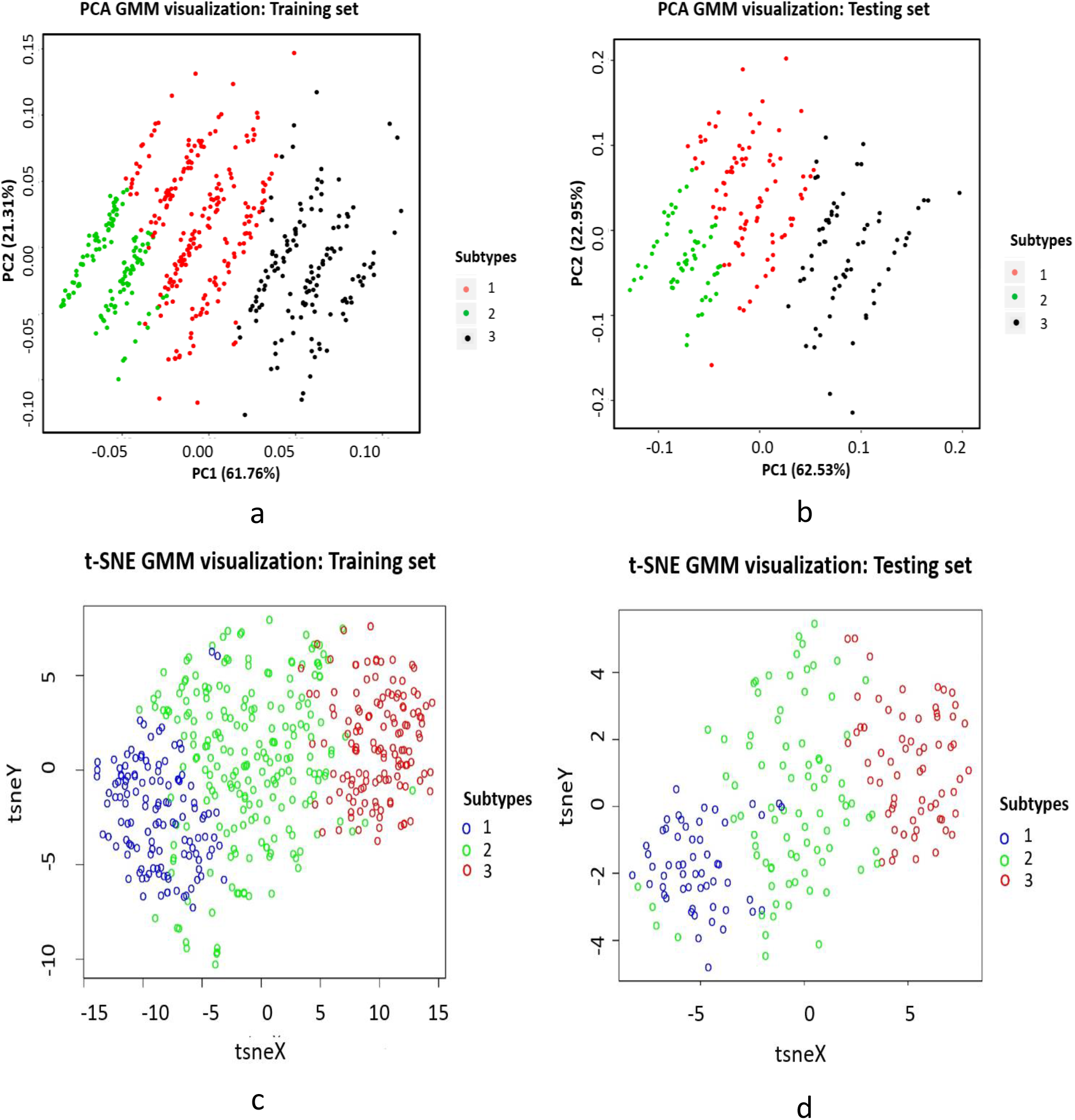
The PCA (a and b) and t-SNE (c and d) visualization of the predicted latent class on the training (A and C) and the testing (B and D) sets.

Table 3, Table 4, and Table 5 depict the descriptive statistics of the explored features stratified by subtypes for all subjects, for just converters, and for just stable subjects, respectively. Notably, most of the features derived from cognitive tests were found to have significantly different means between the medium- and high-risk groups, but that same difference was not found in features from other sources of information. Regarding stable subjects, only the years of education, APOE4 status, and middle temporal gyral volume were not found significantly different between the three subtypes in the testing set. Finally, Figure 4 shows the Kaplan-Meier plots of all three MCI subtypes on the training and testing sets. These plots show the time to conversion, and from them is clear that the low-risk subtype shows a very different trajectory than the at-risk (medium- and high-risk) subtypes. Although the survival curve does not seem very different between subtypes 2 and 3, the log-rank test indicated that they are statistically different (p< 0.05). The survival analyses performed on subjects who progressed to AD indicated that the rate of MCI to AD conversion in at-risk groups was significantly greater than that of the low-risk group in both sets.

**Table 3.** The test results of three subtypes for all MCI subjects. The Mean Standard Error (M±SE) was computed for all features in each subtype. The features were ordered based on the p-value of ANOVA test between the subtypes in the testing set. (* training-testing are statistical different p<0.05)

| MCI Subjects |  |  |  |  |
| --- | --- | --- | --- | --- |
| Testing set (N = 205) |  |  |  |  |
| Features | Subtype 1<br>n=55<br>“Low Risk” | Subtype 2<br>n=85<br>“Medium Risk” | Subtype 3<br>n=65<br>“High Risk” | Anova<br>p-value |
| MMSE | 29.09 ± 0.1* | 28.28 ± 0.11 | 25.17 ± 0.12 | P < 0.001 |
| ADAS13 | 9.87 ± 0.5 | 17.32 ± 0.52 | 21.7 ± 0.66 | P < 0.001 |
| ADAS11 | 6.12 ± 0.35 | 10.69 ± 0.36 | 13.55 ± 0.5 | P < 0.001 |
| RAVLT immediate | 44.89 ± 1.39 | 31.75 ± 0.82 | 27.89 ± 0.9 | P < 0.001 |
| RAVLT learning | 6.02 ± 0.35 | 3.96 ± 0.27 | 2.28 ± 0.22* | P < 0.001 |
| RAVLT % forgetting | 29.8 ± 3.79 | 66.87 ± 3.14 | 75.56 ± 3.29 | P < 0.001 |
| Hippocampus | 7584.1 ± 93.21 | 6453.4 ± 103.65 | 6293.31 ± 137.53 | P < 0.001 |
| Entorhinal | 3861.22 ± 55.96 | 3323.71 ± 70.38 | 3165.85 ± 94.14 | P < 0.001 |
| Ventricles | 26425.07 ± 1643.72 | 44733.4 ± 2416.5 | 49653.96 ± 3438.5 | P < 0.001 |
| FAQ | 0.8 ± 0.17 | 3.93 ± 0.45 | 4.43 ± 0.58 | P < 0.001 |
| CDRSB | 1.04 ± 0.08 | 1.65 ± 0.1 | 1.77 ± 0.11 | P < 0.001 |
| AGE | 69 ± 0.79 | 74.23 ± 0.77 | 74.55 ± 1.05 | P < 0.001 |
| Middle temporal gyrus | 20828.85 ± 289.78 | 19330.93 ± 265.65 | 18954.92 ± 389.74 | P < 0.001 |
| Gender | Male :: 36 *<br>Female :: 19 | Male :: 27<br>Female :: 58 | Male :: 22<br>Female :: 43 | P < 0.001 |
| ICV | 1476558 ± 16799.05 | 1558232 ± 18155.3 | 1555429 ± 22549.6 | P < 0.001 |
| RAVLT forgetting | 3.07 ± 0.37 | 5.18 ± 0.27 | 4.52 ± 0.2 | P < 0.01 |
| APOE4 | 35 / 17 / 3 | 45 / 25 / 15 | 32 / 24 / 9 | 0.22 |
| Education | 16.04 ± 0.39 | 16.29 ± 0.31 | 15.58 ± 0.37 | 0.36 |
| Training set (N = 476) |  |  |  |  |
| Features | Subtype 1<br>n=126<br>“Low Risk” | Subtype 2<br>n=215<br>“Medium Risk” | Subtype 3<br>n=135<br>“High Risk” | Anova<br>p-value |
| MMSE | 29.4 ± 0.06 | 28.08 ± 0.07 | 25.27 ± 0.07 | P < 0.001 |
| ADAS13 | 10.22 ± 0.36 | 16.57 ± 0.38 | 21.33 ± 0.53 | P < 0.001 |
| ADAS11 | 6.46 ± 0.25 | 10.17 ± 0.27 | 13.26 ± 0.39 | P < 0.001 |
| RAVLT immediate | 43.84 ± 0.89 | 32.98 ± 0.6 | 27.54 ± 0.63 | P < 0.001 |
| RAVLT learning | 5.92 ± 0.21 | 4.04 ± 0.16 | 2.89 ± 0.17 | P < 0.001 |
| RAVLT % forgetting | 37.1 ± 2.23 | 66.34 ± 2.12 | 74.91 ± 2.37 | P < 0.001 |
| Hippocampus | 7721.1±77.26 | 6613.1 ± 73.1 | 6196.2 ±86.37 | P < 0.001 |
| Entorhinal | 3983.7±51.64 | 3440.92± 51.99 | 3233.3 ±63.44 | P < 0.001 |
| Ventricles | 27891.21 ± 1280.16 | 42789.54 ± 1549.58 | 42954.19 ± 2134.25 | P < 0.001 |
| FAQ | 1.15 ± 0.16 | 3.83 ± 0.31 | 4.23 ± 0.39 | P < 0.001 |
| CDRSB | 1.12 ± 0.06 | 1.6 ± 0.06 | 1.76 ± 0.08 | P < 0.001 |
| AGE | 68.51 ± 0.6 | 73.14 ± 0.5 | 74.32 ± 0.58 | P < 0.001 |
| Middle temporal gyrus | 21223.97 ± 220.59 | 19632.87 ± 201.19 | 18132.73 ± 255.02 | P < 0.001 |
| Gender | Male :: 60<br>Female :: 66 | Male :: 75<br>Female :: 140 | Male :: 60<br>Female :: 75 | P < 0.001 |
| ICV | 1516545 ± 11587.63 | 1557372 ± 11297.12 | 1540083 ± 15688.69 | 0.26 |
| RAVLT forgetting | 3.88 ± 0.22 | 5.12 ± 0.17 | 4.86 ± 0.18 | P < 0.01 |
| APOE4 | 77 / 45 / 4 | 96 / 90 / 29 | 52 / 66 / 17 | 0.04 |
| Education | 16.69 ± 0.2 | 15.94 ± 0.18 | 14.78 ± 0.29 | P < 0.001 |

**Table 4.** The test results of three subtypes for MCI subjects converted to AD. The Mean Standard Error (M±SE) was computed for all features in each subtype. The features were ordered based on the p-value of t-test that computed between subtypes 2 and 3 in the Testing set. All features were used for clustering except the feature that was shown with *. The Conversion time to AD was computed for evaluation of clustering. (*< 0.05, **<0.01, ***<0.001: p-value)

| Features | MCI to AD conversion |  |  |  |
| --- | --- | --- | --- | --- |
|  | Testing set (N = 74) |  | Training set (N = 170) |  |
|  | Subtype 2<br>n=34<br>"Medium Risk" | Subtype 3<br>n=35<br>"High Risk" | Subtype 2<br>n=86<br>"Medium Risk" | Subtype 3<br>n=78<br>"High Risk" |
| Conversion Time To AD * | 2.51 ± 0.29 | 1.96 ± 0.24 | 2.36 ± 0.19 | 1.93 ± 0.16 |
| MMSE | 28.26 ± 0.18 | 25.29 ± 0.17*** | 28.14 ± 0.1 | 25.31 ± 0.1 |
| ADAS13 | 18.29 ± 0.89 | 23.5 ± 0.86*** | 19.35 ± 0.58 | 23.42 ± 0.66 |
| RAVLT learning | 4.18 ± 0.44 | 2.14 ± 0.29*** | 3.34 ± 0.27 | 2.6 ± 0.21 |
| ADAS11 | 11.65 ± 0.65 | 14.81 ± 0.68** | 11.91 ± 0.43 | 14.44 ± 0.52 |
| RAVLT immediate | 32 ± 1.41 | 27.31 ± 1.12** | 29.58 ± 0.77 | 25.74 ± 0.72 |
| Entorhinal | 3164.6 ± 113.16 | 2877.2 ± 113.92 | 3220 ± 81.27 | 3096.42 ± 85.34 |
| RAVLT forgetting | 5.59 ± 0.4 | 4.74 ± 0.29 | 5.14 ± 0.24 | 4.74 ± 0.24 |
| Hippocampus | 6294.2 ± 150.42 | 5989.1 ± 175.01 | 6269.64 ± 115.22 | 5983.38 ± 104.9 |
| CDRSB | 2.13 ± 0.16 | 2.13 ± 0.14 | 1.91 ± 0.9 | 1.96 ± 0.1 |
| RAVLT % forgetting | 71.66 ± 4.9 | 80 ± 4.47 | 74.38 ± 3.08 | 78.54 ± 3.16 |
| Middle temporal gyrus | 18622.15 ± 399.38 | 17828.26 ± 481.82 | 18893.5 ± 332.22 | 17412.73 ± 308.76 |
| Education | 16.68 ± 0.49 | 15.89 ± 0.44 | 16.05 ± 0.27 | 14.99 ± 0.35 |
| APOE4 | 9 / 16 / 9 | 15 / 14 / 6 | 21 / 51 / 14 | 30 / 35 / 13 |
| AGE | 73.03 ± 1.12 | 74.65 ± 1.35 | 72.74 ± 0.75 | 73.42 ± 0.8 |
| FAQ | 4.91 ± 0.7 | 5.74 ± 0.86 | 5.91 ± 0.56 | 5.31 ± 0.55 |
| Gender | Male :: 11<br>Female :: 23 | Male :: 14<br>Female :: 21 | Male :: 30<br>Female :: 56 | Male :: 40<br>Female :: 38 |
| ICV | 1569399 ± 30124.35 | 1555429 ± 31363.65 | 1580039 ± 17781.46 | 1529759 ± 20933.13 |
| Ventricles | 44233.62 ± 3600 | 44098.4 ± 3604.12 | 45042.6 ± 2557.37 | 43284.28 ± 2516.6 |

**Table 5.** The test results of three subtypes for MCI stable subjects. The Mean Standard Error (M±SE) was computed for all features in each subtype. The features were ordered based on the p-value of ANOVA test between the subtypes on the testing set. (* training-testing are statistical different p<0.05)

| MCI Stable subjects |  |  |  |  |
| --- | --- | --- | --- | --- |
| Testing set (N = 121) |  |  |  |  |
| Features | Subtype 1<br>n=41<br>“Low Risk” | Subtype 2<br>n=51<br>“Medium Risk” | Subtype 3<br>n=29<br>“High Risk” | Anova<br>p-value |
| MMSE | 29.15 ± 0.12 | 28.29 ± 0.14 | 25.03 ± 0.18 | P < 0.001 |
| ADAS13 | 9.69 ± 0.55 | 16.67 ± 0.63* | 19.74 ± 0.92 | P < 0.001 |
| ADAS11 | 5.91 ± 0.39 | 10.06 ± 0.39* | 12.15 ± 0.67 | P < 0.001 |
| RAVLT immediate | 44.51 ± 1.57 | 31.59 ± 1* | 28.14 ± 1.45 | P < 0.001 |
| RAVLT learning | 5.98 ± 0.34 | 3.82 ± 0.35 | 2.45 ± 0.35 | P < 0.001 |
| RAVLT % forgetting | 28.68 ± 4.49 | 63.69 ± 4.05 | 70.5 ± 4.93 | P < 0.001 |
| Ventricles | 26144±1737.6 | 45066.59±3262.3 | 57324.2 ± 6094.1 | P < 0.001 |
| Hippocampus | 7543.71 ±119.04 | 6559.51 ± 139.86 | 6563.28 ± 188.74 | P < 0.001 |
| ICV | 1461997±18424.5<br>* | 1550773±22822.3 | 1608972±33269.7 | P < 0.001 |
| Gender | Male :: 28 *<br>Female :: 13 | Male :: 16<br>Female :: 35 | Male :: 8<br>Female :: 21 | P < 0.001 |
| AGE | 69.16 ± 0.96 | 75.03 ± 1.03 | 74.93 ± 1.64 | P < 0.001 |
| FAQ | 0.85 ± 0.2 | 3.27 ± 0.58 | 3 ± 0.67 | P < 0.01 |
| Entorhinal | 3831.39 ± 66.44 | 3429.75 ± 87.59 | 3514.86 ± 135.79 | P < 0.05 |
| RAVLT forgetting | 3 ± 0.45 | 4.9 ± 0.36 | 4.21 ± 0.28* | P < 0.05 |
| CDRSB | 1.13 ± 0.06 | 1.4 ± 0.08 | 1.5 ± 0.11 | P < 0.05 |
| Education | 16 ± 0.43 | 16.04 ± 0.41 | 15.07 ± 0.62 | 0.23 |
| Middle temporal gyrus | 20763.27 ± 288.61 | 19803.45 ± 340.96 | 20272.14 ± 565.16 | 0.31 |
| APOE4 | 25 / 14 / 2 | 36 / 9 / 6 | 17 / 9 / 3 | 0.35 |
| Training set (N = 279) |  |  |  |  |
| Features | Subtype 1<br>n=99<br>“Low Risk” | Subtype 2<br>n=124<br>“Medium Risk” | Subtype 3<br>n=56<br>“High Risk” | Anova<br>p-value |
| MMSE | 29.37 ± 0.07 | 28.03 ± 0.09 | 25.23 ± 0.11 | P < 0.001 |
| ADAS13 | 10.45 ± 0.4 | 14.82 ± 0.45 | 18.48 ± 0.73 | P < 0.001 |
| ADAS11 | 6.66 ± 0.29 | 9.05 ± 0.31 | 11.67 ± 0.53 | P < 0.001 |
| RAVLT immediate | 43.8 ± 1 | 35.33 ± 0.82 | 29.91 ± 1.07 | P < 0.001 |
| RAVLT learning | 5.86 ± 0.23 | 4.56 ± 0.2 | 3.27 ± 0.28 | P < 0.001 |
| RAVLT % forgetting | 36.39 ± 2.47 | 61 ± 2.82 | 70.21 ± 3.56 | P < 0.001 |
| Ventricles | 27856.04 ±<br>1345.07 | 41527.98 ±<br>1961.6 | 45098.57 ±<br>3784.6 | P < 0.001 |
| Hippocampus | 7692.84 ± 89.18 | 6833.41 ± 92.58 | 6449.59 ± 134.09 | P < 0.001 |
| ICV | 1513539±13110.6 | 1546945±14897.4 | 1549351±23670.7 | 0.12 |
| Gender | Male :: 20<br>Female :: 36 | Male :: 43<br>Female :: 81 | Male :: 45<br>Female :: 54 | 0.22 |
| AGE | 68.96 ± 0.67 | 73.41 ± 0.67 | 75.77 ± 0.8 | P < 0.001 |
| FAQ | 1.01 ± 0.14 | 2.54 ± 0.31 | 2.8 ± 0.51 | P < 0.001 |
| Entorhinal | 3976.63 ± 59.74 | 3573.42 ± 66.86 | 3408.79 ± 90.37 | P < 0.001 |
| RAVLT forgetting | 3.76 ± 0.23 | 5.13 ± 0.24 | 5.02 ± 0.27 | P < 0.001 |
| CDRSB | 1.13 ± 0.06 | 1.4 ± 0.08 | 1.5 ± 0.11 | P < 0.01 |
| Education | 16.65 ± 0.23 | 15.83 ± 0.25 | 14.52 ± 0.49 | P < 0.001 |
| Middle temporal gyrus | 21144.22 ± 240.44 | 20210.97 ± 249.42 | 19096.18 ± 405.57 | P < 0.001 |
| APOE4 | 64 / 31 / 4 | 72 / 38 / 14 | 22 / 30 / 4 | P < 0.01 |

**Figure 4.**
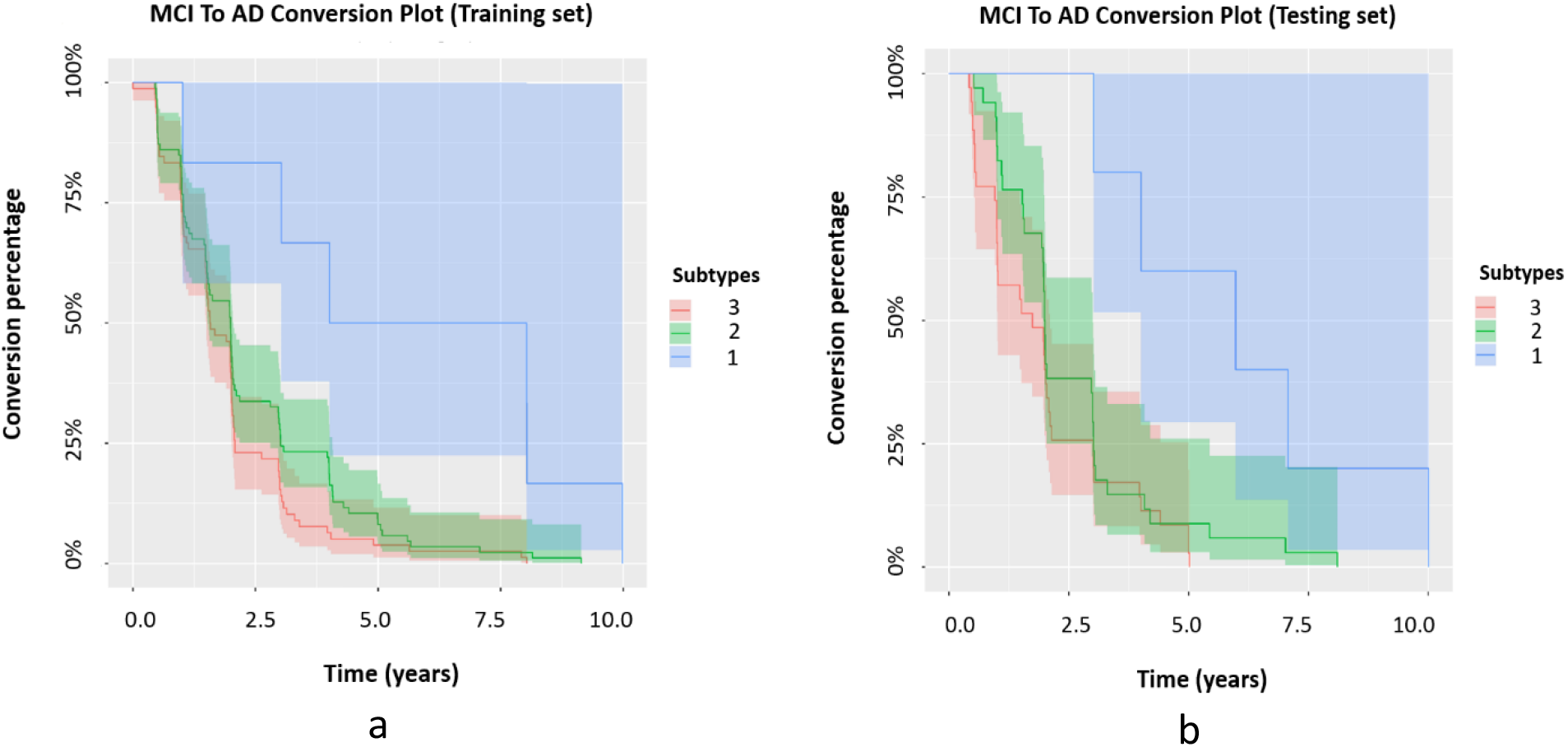
Kaplan-Meier plots for each subtype on a) the Training set and b) the Testing set. There is a significant difference between subtype 1 and subtype 2 (Testing p-value < 0.05) in both plots while subtype 2 and subtype 3 are not significantly different. The log-rank test was used to calculate such differences.

## 4. DISCUSSION

Here, we described three different populations of MCI subjects that were discovered using unsupervised learning via consensus clustering. The discovery of the three MCI subtypes was done using a representative training set of the ADNI database. The modeling of these subtypes with GMM allowed us to predict the MCI classes on an independent testing set. The statistical analysis of the discovered MCI groups identified at least two MCI groups that are at higher risk of progression towards dementia. The OR of converting from MCI to AD for these two groups was very large, namely 8.52 (95%CI 3.22 to 22.56), and slightly larger than the OR between EMCI and LMCI subjects, 7.5 (95%CI 4.9 to 11.5). This implies that the unsupervised clustering successfully discovered the low-risk subjects among the MCI population. Additionally, the two at-risk groups showed different behaviors, having statistically different means for cognitive test-related features, statistically different Kaplan-Meier curves, and contrasting values for the OR of remaining stable and for the OR of converting. The detailed analysis of the at-risk groups indicated that the differences in the two groups were present even after we stratified the analysis for the patients that developed dementia.

Gamberger et al. [23] investigated 5-year longitudinal outcomes and biomarker data from 562 MCI subjects using a novel multilayer clustering algorithm. In that work, two homogenous clusters of MCI subjects with markedly different prognostic cognitive trajectories were identified. A cluster of rapid decliners had a five times larger rate of conversion than that of a cluster of slow decliners. However, some patients did not fit into either cluster, most of them with baseline ADAS scores located between slow and rapid decliners. Our work has classified MCI subjects with the same accuracy, also finding a five times larger rate of conversion between the low-risk and the at-risk groups. But, we assigned all patients to a specific subtype and were able to further subdivide the at-risk group into significantly different subtypes (medium- and high-risk).

The herein presented unsupervised analysis of the ADNI data suggests that subjects at MCI may represent different AD subtypes that can be discovered at the prodromal phase of AD. Also, it may be worth exploring what other differences exist between the two at-risk groups, and these studies may highlight differences in etiology and possible differences in treatment response among these two groups.

The current study is limited by the fact that only ADNI subjects with their corresponding clinical/medical and imaging features were used for this study. Our findings need to be validated in an independent cohort; nevertheless, the robust strategy of using a hold-out sample for inferring the properties and differences among the discovered groups indicates that there are at least three different MCI subtypes in the ADNI cohort.

## 5. CONCLUSION

In this search, we took advantage of consensus data clustering and GMM to discover the latent classes from a set of AD-relevant characteristics of MCI subjects. After identifying MCI subtypes, the association of discovered classes with MCI to AD conversion were described. Our findings revealed three different subtypes of patients with MCI at early stage. At-risk groups showed a different trajectory than the low-risk subtype. Also, the characterization of these subgroups indicated that there are further substantial differences in some features that we considered studying amongst subgroups.

## Data Availability

Data used in the preparation of this study was obtained from the TADPOLE challenge standard data sets. The TADPOLE sets were derived from the Alzheimers Disease Neuroimaging Initiative (ADNI) study (adni.loni.usc.edu).

https://tadpole.grand-challenge.org

